# Synergy-based functional electrical stimulation for gait rehabilitation in chronic stroke: a pilot study

**DOI:** 10.1101/2025.05.21.25328035

**Authors:** Jackson T. Levine, Xin S. Yu, Rebecca Muñoz, Alaina Fiorenza, Tyler Smith, Irina Djuraskovic, JD Peiffer, Emilia Ambrosini, Simona Ferrante, Rebecca Webster, Jon Sakai, Jeremiah Robison, Elliot J Roth, Jozsef Laczko, Ronald J Cotton, Alessandra Pedrocchi, José L. Pons

## Abstract

Chronic stroke gait disorders involve impaired motor coordination. While high-intensity gait training (HIGT) is supported by current clinical practice guidelines, and Functional Electrical Stimulation (FES) to tibialis anterior addresses foot drop, extending FES to multiple muscles may improve functional outcomes. Leveraging a fast-to-don FES sleeve, we tested feasibility and preliminary efficacy of a personalized multichannel FES (MFES) intervention based on the individual’s motor coordination impairment paired with HIGT. Fourteen chronic stroke survivors were randomly assigned to either HIGT or MFES+HIGT for six weeks. Feasibility was evaluated by measuring setup time and collecting feedback from participants and four therapists. Gait speed, endurance, gait biomechanics, and muscle synergies were assessed at baseline, midpoint, post-training, and one-month follow-up. System setup time plateaued at 4.53 minutes by the ninth session. Both participants and therapists rated the intervention highly feasible, acceptable, and usable. Adherence was high, with no dropouts in the MFES+HIGT group. While most participants reached target heart rate zones, those with severe impairments (N=3, <0.4 m/s gait speed) struggled to maintain these levels. Despite the small sample, only the MFES+HIGT group demonstrated significant endurance gains from baseline to post-training and follow-up, while both groups improved walking speed, impaired limb step length, and muscle synergy similarity to normative data. When excluding household ambulators, only the MFES+HIGT group showed post-training and follow-up gains in endurance and self-selected walking speed. This study demonstrates that synergy-based MFES is feasible for integration into chronic stroke gait rehabilitation supports larger-scale trials to validate clinical efficacy and identify responders.

## I. Introduction

GAIT impairment is one of the most common and debilitating consequences of stroke, affecting up to 80% of survivors and significantly limiting mobility and independence.[1] Post-stroke gait is often slower, more fatiguing, and marked by poor coordination and compensatory strategies.[2-4] Although high-intensity gait training (HIGT) is supported by current clinical practice guidelines [5] and has shown substantial benefits during the subacute phase of recovery,[6, 7] recovery potential with current approaches is still modest during the chronic phase.[8] Despite the availability of novel assistive technologies such as robotic exoskeletons and electrical stimulation devices, their clinical use remains limited due to cost, complexity, and lack of integration into existing therapy workflows.[9] As a result, HIGT often follows a generalized approach and fails to fully address the individualized motor coordination deficits that persist in chronic stroke.

When paired with task-oriented movement intent, Functional Electrical Stimulation (FES) can induce peripheral benefits [10] and enhance central nervous system neuroplasticity [11-15] by providing timed muscle activation and sensory feedback and helping to reshape neural control through repetitive training.[16, 17] It is most commonly used to treat foot drop through stimulation of the tibialis anterior (TA) during swing phase, reducing compensatory patterns such as circumduction and hip hiking.[2, 18] However, single-muscle stimulation has demonstrated limited benefits over HIGT, particularly in chronic stroke populations.[19] Extending FES to multiple muscles has shown some moderate improvements, such as improved endurance, gait velocity, and gait symmetry with the addition of the gluteus medius (GM) [20, 21] or reduced energy expenditure with the addition of the gastrocnemius (GAS).[22, 23] The complexity of post-stroke gait, which often includes impaired intermuscular coordination, underscores the need for broader and more individualized stimulation strategies—potentially involving a larger array of muscles. One prominent indicator of this impairment is the reduction in number of muscle synergies and the presence of abnormal coordination patterns, which can be quantified through muscle synergy analysis of electromyography (EMG) signals.[24] Muscle synergies represent the coordinated activation patterns produced by the nervous system to control movement and are linked to biomechanical functions of gait: weight acceptance, knee flexion, propulsion, foot clearance.[24-26] After stroke, these synergies often become disrupted, leading to inappropriate timing of muscle activations and abnormal coactivation of muscles. Notably, individuals with impaired synergies (e.g., reduced number of synergies, reduced similarity of synergies to neurologically intact individuals) exhibit more impaired gait (e.g., slower speed, shorter step length, asymmetries).[27, 28] Those who demonstrate improved locomotor function following rehabilitation often show muscle synergy patterns that more closely resemble those of neurologically intact individuals,[29-31] suggesting that rehabilitation can restore more physiologically appropriate muscle coordination, thereby improving gait function and biomechanics. These findings also highlight the potential of muscle synergies to serve both as biomarkers and as therapeutic targets for stroke recovery.

To better target these coordination deficits, we propose a personalized rehabilitation intervention that delivers multichannel FES (MFES) based on an individual’s impaired muscle synergies precisely timed during HIGT to reduce gait impairment in chronic stroke survivors. This approach provides feedback to all major leg muscles according to the individual’s specific neural coordination deficits, quantified through muscle synergy analysis at baseline. Our preliminary work [32] and that of colleagues [33] using manually placed electrodes demonstrated proof of concept in two individuals each with chronic stroke during a four-week gait training intervention, and found improved walking speed and similarity of synergies to neurologically intact individuals. Additionally, the FDA-approved Cionic Neural Sleeve provides a clinically viable, quick to don delivery platform for wireless and customizable MFES to the major muscle groups of the leg, which is currently approved for home use in individuals with upper motor neuron injuries.[34, 35]

In this pilot study, we aimed to evaluate clinical feasibility and preliminary efficacy of a six-week synergy-based MFES combined with HIGT protocol compared to HIGT alone in chronic stroke survivors. We hypothesized that therapists would quickly become proficient in using the Neural Sleeve and be able to integrate it efficiently within typical therapy timelines. Furthermore, we hypothesized that participants receiving personalized synergy-based MFES would exhibit greater improvements in gait speed and endurance, supported by changes in biomechanics and muscle coordination, compared to those receiving conventional HIGT alone.

## II. Methods

### A. Device

This study used the Cionic Neural Sleeve (Fig. 1B), an FDA-approved device (510(k) number K21362) intended to provide ankle dorsiflexion and/or plantarflexion in adult individuals with foot drop and/or to assist knee flexion or extension in adult individuals with muscle weakness related to upper motor neuron disease/injury (e.g. stroke, damage to pathways to the spinal cord).[34, 35] The sleeve integrates EMG recording and biphasic stimulation electrodes from key lower limb muscles (tibialis anterior (TA), gastrocnemius (GAS), rectus femoris (RF), vastus lateralis (VL), semitendinosus (ST), and biceps femoris (BF)) with two embedded inertial measurement units (IMUs) on the shank and thigh. A custom external hip unit was developed to target the gluteus medius (GM), also equipped with EMG and IMU sensors (depicted in right panel of Fig. 1B). EMG signals were sampled at 2000 Hz and IMU data at 100 Hz. The study protocol was approved by the Northwestern University Institutional Review Board (STU00218244, date of initial approval: 8/17/2023, clinical trial number: NCT06099444) and conducted in accordance with the Declaration of Helsinki.

**Fig. 1.**
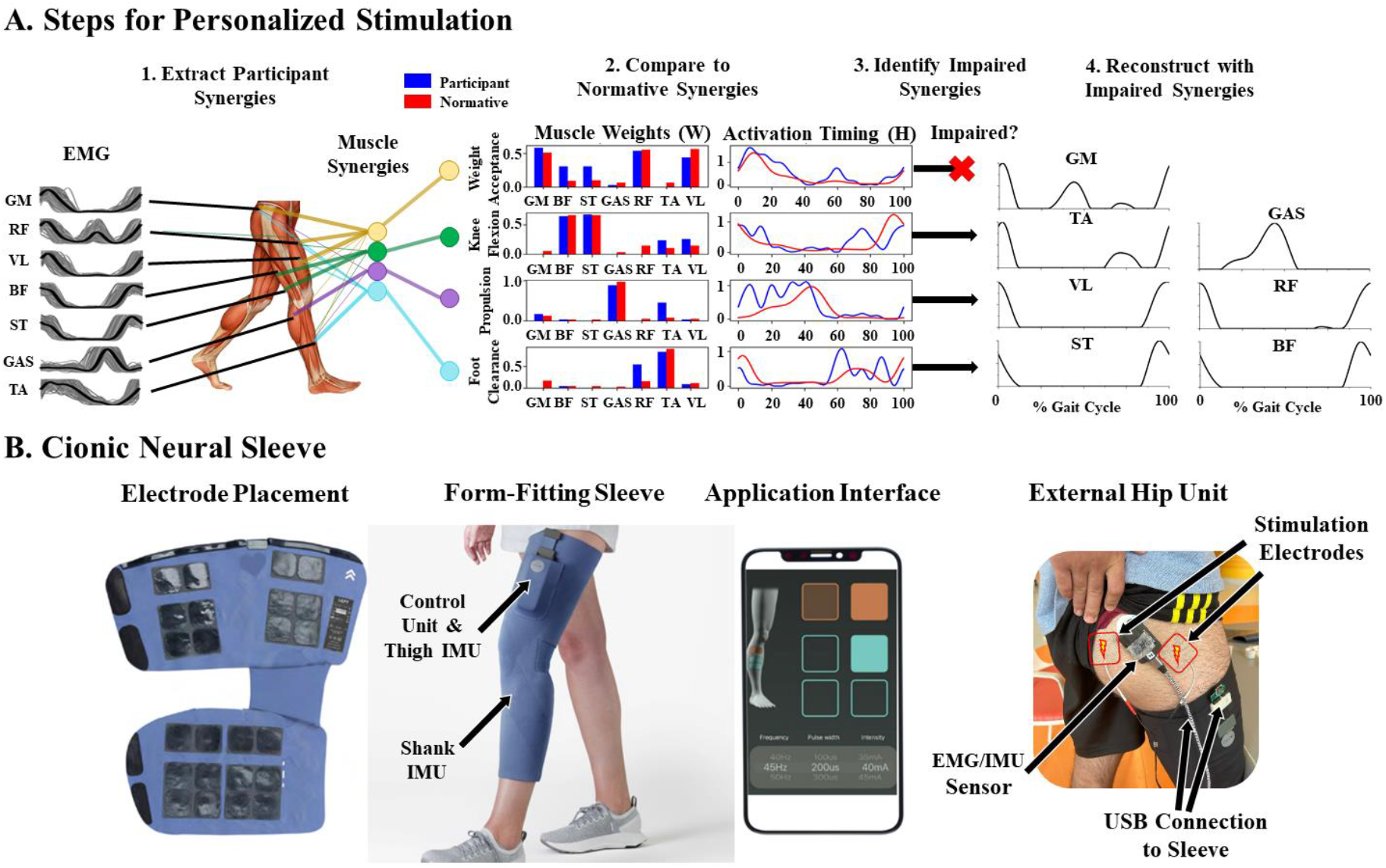
Personalized Synergy-Based Stimulation. A) Steps for identifying impaired synergies, computing personalized stimulation based on the identified impaired synergies, and precisely timing stimulation. B) Cionic Neural Sleeve with electrode placement, sleeve fit, and the application. The placement of the external hip unit is also displayed, with labels for the stimulation electrodes and sensor for recording electromyography (EMG) and inertial measurement unit (IMU) signals. GM = Gluteus Medius, GAS = Gastrocnemius, TA = Tibialis Anterior, VL = Vastus Lateralis, RF = Rectus Femoris, BF = Biceps Femoris, ST = Semitendinosus.

### B. Population

Fourteen individuals with chronic, unilateral stroke (>6 months) and reduced but independent ambulation were recruited. Exclusion criteria included multiple strokes, other neurological or orthopedic impairments, contraindications to FES, and concurrent lower limb studies. Participants were randomly assigned to one of two groups: 1) HIGT or 2) HIGT in combination with synergy-based MFES (MFES+HIGT). All participants provided informed consent at the initial visit and then completed a baseline assessment. Fourteen neurologically intact individuals were recruited to establish normative synergies for comparison with stroke synergies.

To assess clinical feasibility, four physical therapists from Shirley Ryan AbilityLab were recruited to deliver the MFES+HIGT training protocol. All therapists provided informed consent and completed two hours of training on both the rehabilitation protocol and use of the Cionic Neural Sleeve prior to being paired with a post-stroke participant for the six-week intervention. On average, therapists had 4.5 years of experience in post-stroke gait rehabilitation and were familiar with single-channel FES for foot drop (primarily using the Bioness device to stimulate the TA).

### C. Intervention

Participants trained three times weekly for six weeks (eighteen sessions), walking on a treadmill with a harness (no weight support) per clinical guidelines.[5] Each session included a five-minute warm-up and cool down at self-selected velocity (SSV, determined at baseline assessment) and 20 minutes of HIGT with or without synergy-based MFES. Therapists adjusted treadmill speed and incline to challenge participants to reach 70-85% of their maximum heart rate (HR) [5] and Borg Rating of Perceived Exertion (RPE) between 15 and 17.[36] HR was measured using a Polar OH1 sensor over wrist extensors or biceps.[37, 38]

For participants in the MFES+HIGT group, therapists donned the sleeve and external hip unit on the impaired limb. Maximum comfortable stimulation amplitudes (15–100 mA) were determined per muscle, with fixed frequency (30 Hz) and pulse width (300 µs). Personalized stimulation was applied during the 20-min training segment, where parameters could be adjusted as necessary. Stimulation parameters were saved after the first training and updated for each training, if needed. Setup time was recorded to assess feasibility.

### D. Personalized Stimulation Control

Personalized stimulation has been designed as described in our previous work,[32] validating the concept (Fig. 1A).

#### 1) Participant EMG Processing and Muscle Synergies

Participant EMG was recorded at baseline during treadmill walking at SSV (determined during baseline 10MWT). Signals were bandpass filtered (3rd order, 40-400 Hz cutoff, Butterworth filter), rectified, and low pass filtered (3rd order, 5 Hz cutoff, Butterworth). They were segmented into gait cycles, interpolated from 0 to 100% of the gait cycle, and normalized to peak-to-peak amplitude (Fig. 1A Step 1). Normative muscle synergy weights (W) and activation profiles (H) were computed as the average of fourteen neurologically intact individuals (age: 46.9±17.0) walking at ten speeds ranging from 0.45 m/s to 1.8 m/s for one minute each using nonnegative matrix factorization (NNMF) and a variance accounted for threshold of 0.9 to define the number of synergies (four synergies were extracted on average).[39] To ensure accurate comparison between participant and normative synergies—and to prevent incorrect matching between non-corresponding components—participant-specific synergy weights and activation profiles were computed using an iterative non-negative matrix reconstruction approach where normative weights were fixed to reconstruct the participant activation profiles and the normative activation profiles were fixed to reconstruct the participant weights; full algorithmic details are provided in Ferrante et al.[32]

#### 2) Identification of Impaired Participant Synergies

Participant synergies were compared to average normative synergies, and the degree of deviation was quantified using cosine similarity of weights W, circular cross correlation coefficient of activation profiles H, and circular cross correlation time lag of activation profiles H.[32] The time lag was computed as 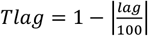, where lag represents a percentage of the gait cycle (value between -50 and 50%). The mean and standard deviation for all three metrics across muscle synergies of neurologically intact individuals were computed in comparison to the average normative synergies. For each participant synergy, comparisons to normative synergies were computed, and a synergy was classified as impaired if any of the three metrics deviated by more than two standard deviations from the normative (Fig. 1A Step 3).

#### 3) Personalized Participant Stimulation Profiles

Only impaired synergies were used to drive the personalized stimulation profile development. The normative weights and activation profiles of synergies that were deemed impaired for each participant were multiplied to define a stimulation profile for each muscle. The stimulation profile was normalized between zero and one. To prevent continuous stimulation throughout the gait cycle and minimize fatigue, only stimulation values exceeding 0.2 were included in the stimulation profile. These values were then scaled to the maximum stimulation amplitude set by the therapist through the interface (Fig. 1A Step 4). Stimulation profiles were precisely triggered by the IMU sensors in the shank and thigh.

### E. Assessment Visits and Outcome Measures

#### 1) Feasibility

To obtain participant feedback on the experimental intervention, participants in both groups completed the Patient Perceived Usefulness Questionnaire, with the prompt “With respect to your walking ability, how would you describe yourself now compared to before starting your rehabilitation program?” at the follow-up assessment via a scale from -5 (Much Worse) to 5 (Completely Recovered).[40] Adverse reactions were recorded.

To assess clinical feasibility, time required to don the sleeve and set stimulation parameters was recorded for each session for the first participant each therapist trained. To assess the ability of therapists to challenge their participants to achieve the requirements of HIGT, mean HR, peak HR, percentage of time spent in the target HR zone, and RPE were evaluated for each session. Therapists also completed the Therapist Perceived Usefulness Questionnaire, with the prompt “With respect to the rehabilitation program used for this patient, how likely would you be to integrate this into your clinical practice?”, ranging from -5 (Very Unlikely) to 5 (Very Likely). [40] Additionally, therapists completed two more surveys: Brooke System Usability Scale [41] and Weiner Acceptability, Appropriateness and Feasibility of Intervention Measure.[42] All therapist forms were collected after completing at least six training sessions.

#### 2) Assessment Visit

Assessments were performed at baseline, midpoint, post, and one month after completing the training. The baseline assessment was completed one week prior to the beginning of training. During this week, the participant wore a step-counting device (ActiGraph, USA) [43-45] to track average daily number of steps taken as a measure of participation. This was repeated immediately following completion of all training sessions. At each assessment, participants completed the 10-Meter Walk Test (10MWT) at both SSV and fast velocity (FV) and the 6-Minute Walk Test (6MWT). During the 10MWT at SSV, participants were recorded using a markerless motion capture system.[46] Participants were permitted to use the same assistive device for all assessment points (e.g., cane or walker), but ankle-foot orthoses were not allowed to assess intrinsic ankle motor control impairments. Three levels of ambulation are reported based on the self-selected walking speed during the 10MWT at baseline: household (<0.4 m/s), limited community (0.4-0.8 m/s), and community (>0.8 m/s).[47] Participants then donned the Neural Sleeve and external hip sensor on the impaired limb. Following a two-minute warm-up, participants walked on a treadmill for two minutes at their SSV while IMU angular data and EMG from the same muscles used for stimulation were recorded.

#### 3) Gait Biomechanics

Markerless motion capture data taken during 10MWT SSV was processed according to Cotton et al.[46, 48] Videos were collected from multiple calibrated and synchronized cameras. Participant tracking using EasyMocap [49] and 2D joint location estimation using MeTRAbs-ACAE [50] was facilitated using PosePipe.[51] We used an end-to-end reconstruction approach to jointly learn the body scale parameters and joint angles of a differentiable biomechanical model most consistent with video observations.[48] From these biomechanical reconstructions, we extracted gait event timings [52, 53] and calculated spatiotemporal measures: impaired limb step length, step width, double support percent, and impaired limb single support percent. Kinematic analysis focused on joint movements and phases specific to stroke impairment on the impaired limb: peak ankle dorsiflexion during swing (foot clearance), peak hip abduction during swing (circumductory gait compensation), peak hip extension during stance (propulsion), and peak knee extension during stance (stance stability and hyperextension).

#### 4) Muscle Synergies

Cosine similarity and circular cross correlation were assessed for the impaired limb compared to normative synergies. These measures were computed for each synergy, and the average among synergies impaired at baseline was reported. Additionally, the number of synergies extracted from NNMF where variance accounted for plateaued was reported.[54]

### F. Statistics

SPSS 26.0.0.1 was used for statistical analysis. To compare group demographics, a linear mixed model was used with Group (HIGT, MFES+HIGT) as a fixed effect and Participant as a random effect. HR and RPE were compared using a linear mixed model with Session Number and Group as fixed effects and Participant and Therapist as random effects. A linear plateau model was applied in RStudio 2025.05.0 to determine the inflection point at which setup time plateaued. Setup durations from 57 total sessions conducted by four physical therapists (PT1: 13 sessions, PT2: 15 sessions, PT3: 15 sessions, PT4: 14 sessions) were analyzed.

For efficacy outcome measures, a linear mixed model was used with Group and Visit (pre, mid, post, follow-up) as fixed effects and Participant as a random effect. Assumptions of linearity, independence, homoscedasticity, normality of residuals, and absence of multicollinearity were verified and transformations applied where needed according to Shapiro-Wilk’s test.[55, 56]. Non-normal data were analyzed using generalized linear mixed models. Post-hoc comparisons were made with Bonferroni corrections and α = 0.05.

Due to the inability of household ambulators to reach target HR, an exploratory sub-group analysis was conducted on the primary outcomes – clinical assessments – excluding participants that were categorized as household ambulators at baseline (i.e., self-selected walking speed < 0.4 m/s, N=2 from MFES+HIGT, N=1 from HIGT) using a linear mixed model with Group and Visit as fixed effects with Participant as a random effect.

## III. Results

### A. Participant Demographics

Of eighteen screened, fourteen participants (three females per group) completed training. Three did not pass screening while one in the HIGT group dropped out of the study due to pre-existing medical issues before mid-point and therefore was not included in the analysis. Demographics are displayed in Table I. There were no differences between groups in age (HIGT: 60.6±10.7, MFES+HIGT: 57.7±10.2, p=0.331), years since stroke (p=0.703), baseline 10MWT SSV (p=0.625), nor baseline 6MWT (p=0.462).

**TABLE 1.**
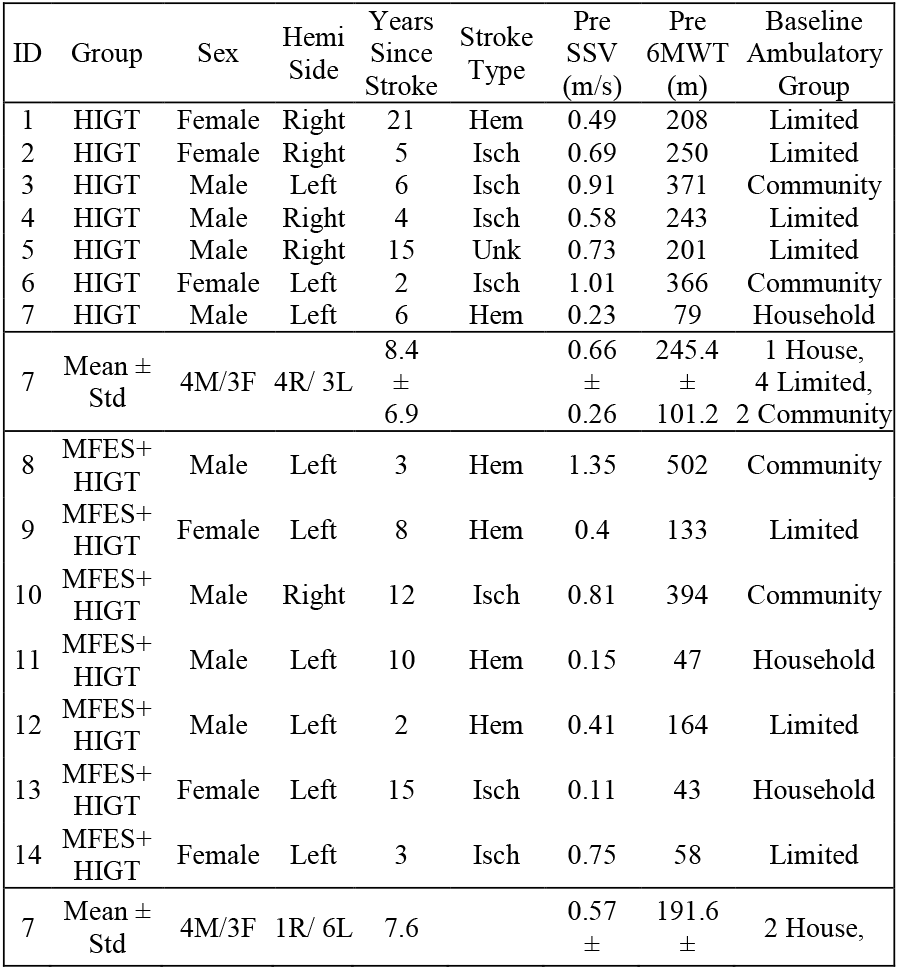
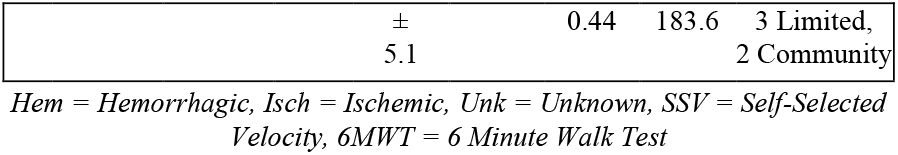
Participant Demographics

### B. Training and Feasibility

#### 1) Training Measures

Training and cardiovascular intensity results demonstrate that on average both groups maintained target HR intensities for 50% of each session, consistent with other HIGT studies.[7] In addition, average RPE was 14.1±3.2 for HIGT and 14.9±2.9 for MFES+HIGT, indicating that participants perceived the intensity of the intervention close to ‘Hard’. The percentage of time spent in the target heart rate zone was 72.6±30.2% and 49.2±37.6% for HIGT and MFES+HIGT, respectively. The three household ambulators (2 in MFES+HIGT, 1 in HIGT) spent on average 22.9±14.3% in the target heart rate zone, and when excluded from the analysis, participants in the HIGT group averaged 78.1±21.1% and MFES+HIGT group averaged 62.9±29.6%. Despite the lower percentage of time spent in the target heart rate zone, these participants averaged above 13 for their RPE rating. Regardless, there was no main effect of group, session, nor group^*^session interaction for mean HR, peak HR, percentage of time in HR zone, nor mean RPE. P-values are reported along with mean and standard deviations for all metrics considering all participants and all sessions (see Table II). There were no adverse reactions in either group to training.

**TABLE 2.** Training and Cardiovascular Intensity Details

| Metric | HIGT | MFES+HIGT | P-value |
| --- | --- | --- | --- |
| Mean HR (% Max) | 75.0 % (69.0 % - 79.0 %) | 67.0 % (65.0 % - 81.0 %) | 0.31 |
| Peak HR (% Max) | 84.0 % (75.0 % - 85.0 %) | 74.0 % (73.0 % - 89.0 %) | 0.28 |
| Percent Time in HR Zone (%) | 74.0 % (42.0 % - 96.2 %) | 40.5 % (21.8 % - 93.0 %) | 0.17 |
| Mean RPE | 14.6 (12.0 - 16.0) | 16.0 (13.2 - 16.6) | 0.65 |

Median and interquartile range are reported. A linear mixed effects model with fixed effects of group and session and a random effect of subject was conducted, and the p-value of the group effect is reported. There was also no effect of session nor a group\*session interaction.
HIGT = High-Intensity Gait Training, MFES = Multichannel Functional Electrical Stimulation, HR = Heart Rate,
RPE = Borg Rating of Perceived Exertion (Range = 6-20).

#### 2) Clinical Feasibility

System setup time for the four therapists was recorded with four of the seven participants in the MFES+HIGT group and assessed for feasibility of clinical implementation, (see Fig. 2). Each data point represents a single session with a single participant for each therapist. The inflection point was identified at 8.48 sessions and the slope of the learning phase was -0.758 minutes per session. The curve plateaued at 4.53 minutes. Therapists also completed the Therapist Perceived Usefulness Questionnaire on whether they would implement synergy-based MFES+HIGT into their own clinical practice, and reported a mean score of +3.6±0.48. Therapist feedback highlighted ease of use to don the sleeve and update stimulation parameters throughout training. For the Implementation Outcome Measures,[42] which range from 1 to 5 and measure feasibility, appropriateness, and acceptability of intervention, therapists reported an average score of 3.9±0.13, 4.1±0.13, and 4.0±0.00, respectively. Lastly, on the 10-item System Usability Scale, average therapist score for synergy-based MFES was 72.15±7.5 indicating an acceptable rating.[57]

**Fig. 2:**
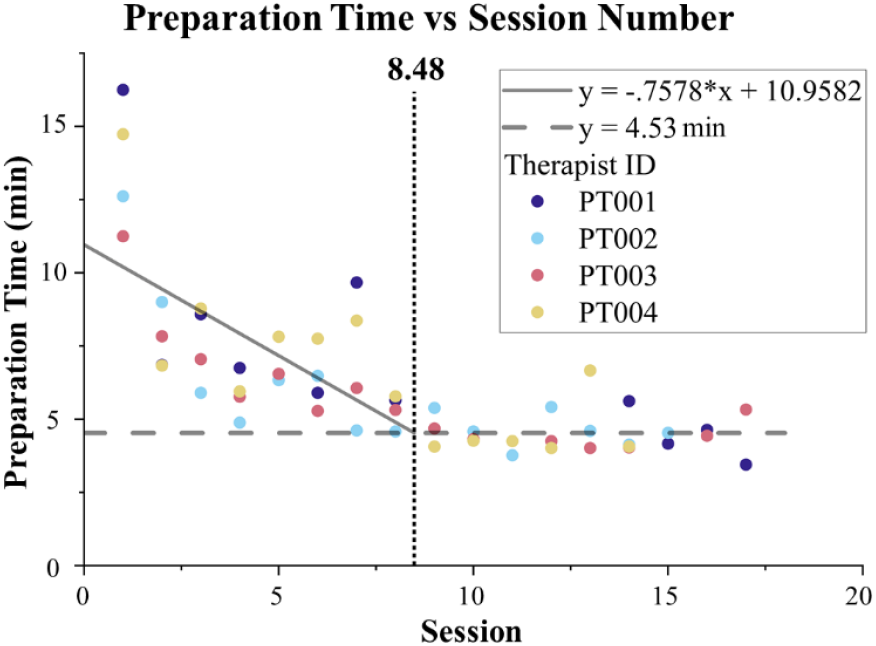
Preparation Time vs Number of Sessions. The time to don the sleeve and set parameters was recorded for each session for each of the four therapists, who are each depicted with a different color as specified in the legend. An inflection point analysis was conducted to identify the learning region and the plateau region. Each data point represents a single session for each therapist with the participant with whom they were paired.

We also collected participant feedback after completing the training with the Patient Perceived Usefulness Questionnaire. On average, the MFES+HIGT group rating was +3.43±1.51, while the HIGT group rating was +3.14±0.38, which were not significantly different (p = 0.636). Qualitative feedback from both groups highlighted improved endurance, but notably, multiple participants in the MFES+HIGT group emphasized reduced reliance on assistive devices.

### C. Training Efficacy

No differences were identified between groups at baseline for clinical, biomechanical, nor synergy metrics (all p>0.05). All data are displayed in Supplemental Table I.

#### 1) Clinical Assessments

Clinical assessments are displayed in Fig. 3 and Supplemental Table I. For the 10MWT SSV, there was an effect of visit (p=0.001) but no effect of group (p=0.705) nor interaction (p=0.735). Post-hoc analysis of visit revealed improvements at midpoint (p=0.048), post (p=0.001), and follow-up (p=0.005) compared to pre. Two out of seven participants achieved improvements greater than minimal clinically important difference (MCID, 0.14 m/s [58]) in the MFES+HIGT group and only one in the HIGT group. Similarly, the 10MWT FV exhibited an effect of visit (p=0.021) but no effect of group (p=0.392) nor interaction (p=0.638). Post-hoc analysis of visit revealed an improvement at post (p=0.015) compared to baseline, but not for pre to mid (p=0.075) or pre to follow-up (p=0.063). For the 6MWT, there was an effect of visit (p=0.011) and an interaction (p=0.026), but no effect of group (p=0.990). Post-hoc analysis of the interaction revealed that, in the MFES+HIGT group only, there was an improvement from pre to post (p=0.002) and pre to follow-up (p=0.009). Four out of seven participants reached MCID of the 6MWT (50 m [58]) in the MFES+HIGT group while only one achieved MCID in the HIGT group. Finally, average number of steps per day did not exhibit any effects (all p>0.406, see Supplemental Table I).

**Fig. 3:**
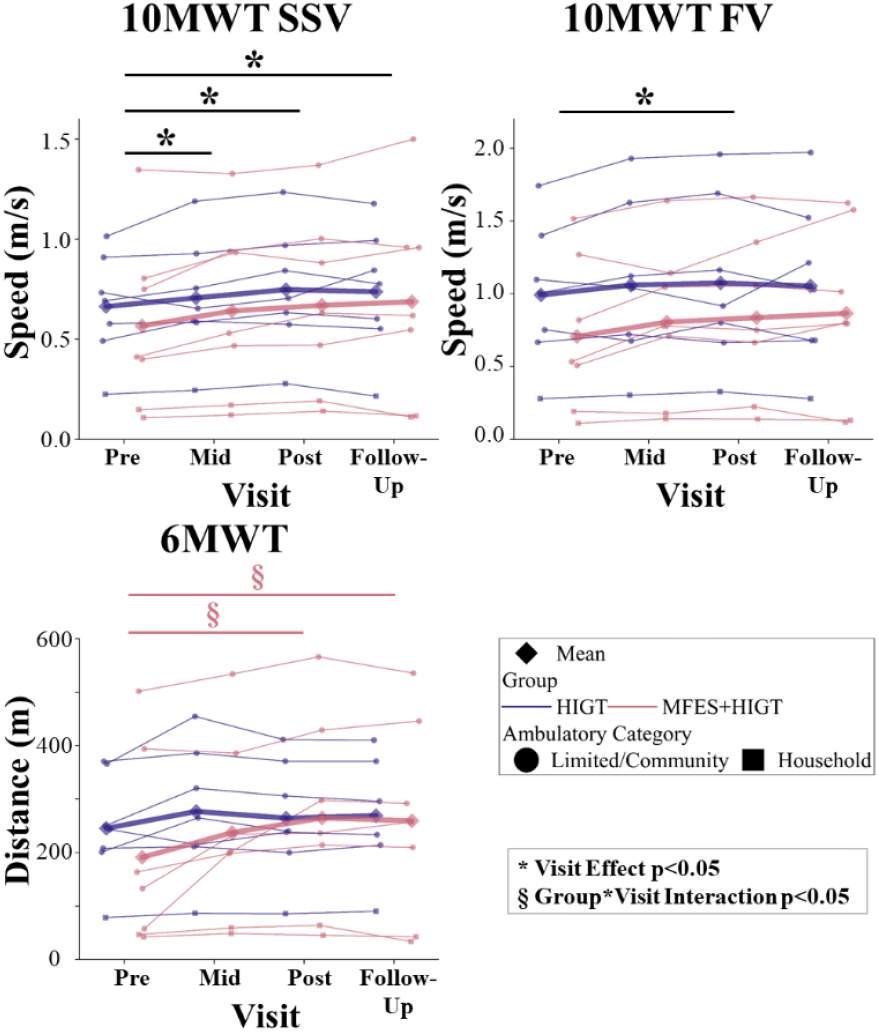
Clinical Assessments. The 10 Meter Walk Test (10MWT) at self-selected velocity (SSV) and fast velocity (FV) and the 6 Minute Walk Test (6MWT) are presented. Improvements for all metrics are positive. Individual data points for each participant are displayed with household ambulators as squares and limited and community ambulators as circles. Significant effects of visit are depicted by black lines and ^*^ where p<0.05. Significant group^*^visit interactions are depicted by lines matching the color of the group associated with the interaction and § where p<0.05.

#### 2) Gait Biomechanics

Selected biomechanics are displayed in Fig. 4 and all data is displayed in Supplemental Table I. Impaired limb step length exhibited an effect of visit (p=0.007), while group (p=0.434) and the interaction (p=0.922) did not reach significance. An improvement (increase) was detected from pre to follow-up (p=0.045) and midpoint to follow-up (p=0.047), while pre to post did not reach significance (p=0.088). Similarly, step width exhibited an effect of visit (p=0.033), while group (p=0.414) and the interaction (p=0.110) failed to reach significance. However, there were no significant contrasts. Neither impaired limb single support nor double support exhibited effects of group, visit, nor interaction (all p>0.099).

**Fig. 4:**
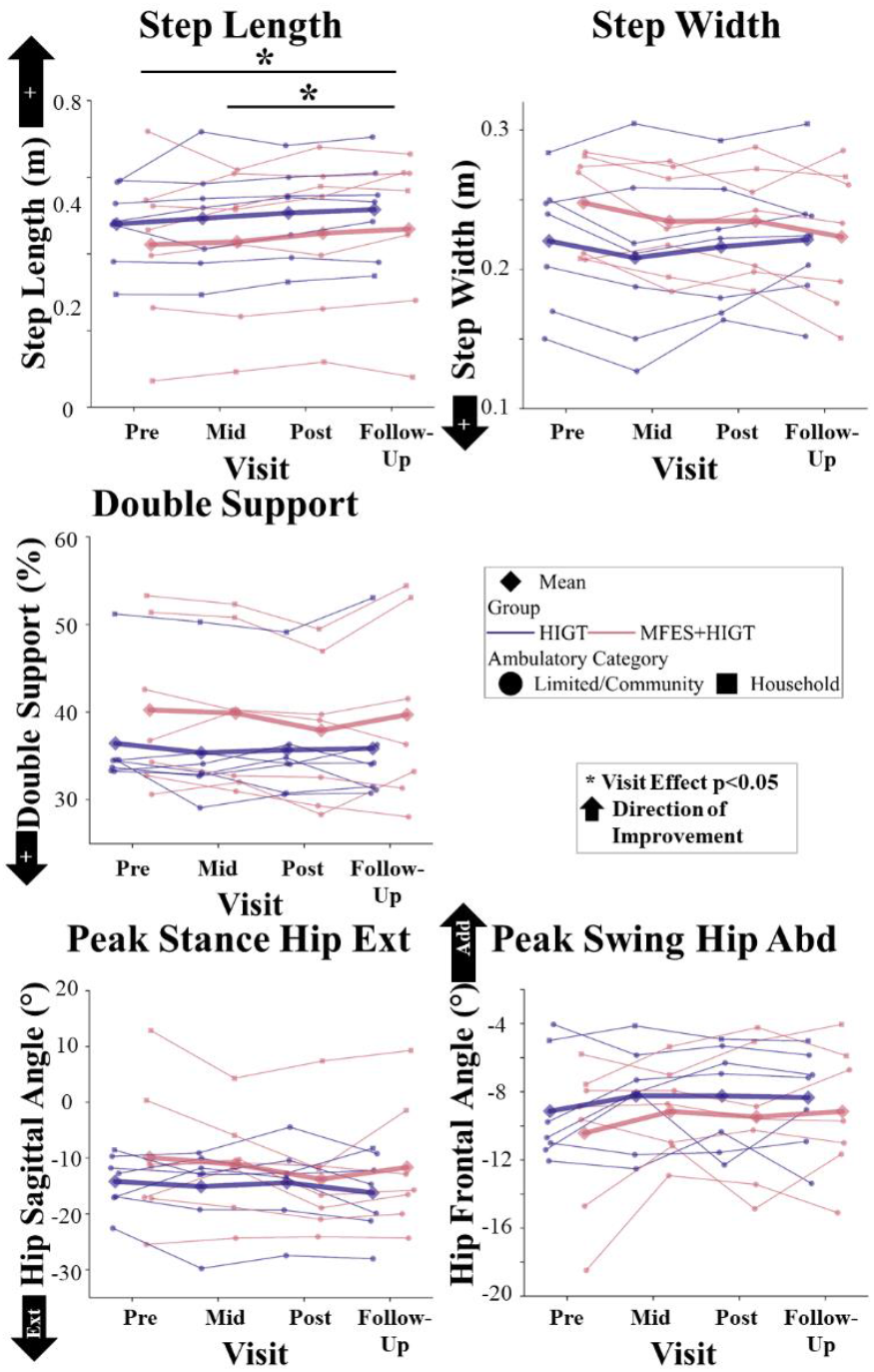
Biomechanics. Top panel: impaired limb step length, step width, and double support. Bottom panel: peak impaired limb hip extension angle during stance, peak impaired limb hip abduction (Abd) angle during swing. Extension (Ext) for hip sagittal angle is negative. Adduction (add) for hip frontal angle is positive. Arrows are displayed to depict the direction of improvement. Individual data points for each participant are displayed with household ambulators as squares and limited and community ambulators as circles. Significant effects of visit are depicted by black lines and ^*^ where p<0.05.

Ankle dorsiflexion during swing, knee extension during stance, and hip abduction during swing also did not exhibit any effects of group, visit, nor interactions (p>0.055). Hip extension during stance phase exhibited an effect of visit (p=0.040), but no contrasts achieved significance (p>0.088) nor did the effect of group (p=0.479) and interaction (p=0.216). In general, there was a modest trend toward increased extension.

#### 3) Muscle Synergies

On average, four muscle synergies were extracted from neurologically intact individuals, and therefore, four synergies were used for the average normative data. Muscle synergies of participants were compared with average normative data and results are displayed in Fig. 5 and Supplemental Table I. On average, 2.1±1.6 and 2.3±0.8 synergies were impaired at baseline for the HIGT group and MFES+HIGT group, respectively. In both groups, the synergy related to propulsion was most frequently impaired (HIGT: 5/7, MFES+HIGT: 6/7) while the synergy related to weight acceptance was least frequently impaired (2/7 in both groups). Cosine similarity had no effect of group (p=0.253), visit (p=0.244), nor interaction (p=0.427). Circular cross correlation coefficient had an effect of visit (p=0.005) but no effect of group (p=0.473) nor interaction (p=0.850). Post-hoc analysis revealed that correlation coefficient improved from pre to post (p=0.024) and pre to follow-up (p=0.049). The number of components extracted from NNMF had no effect of group (p=0.150), visit (p=0.095), nor interaction (p=0.771).

**Fig. 5:**
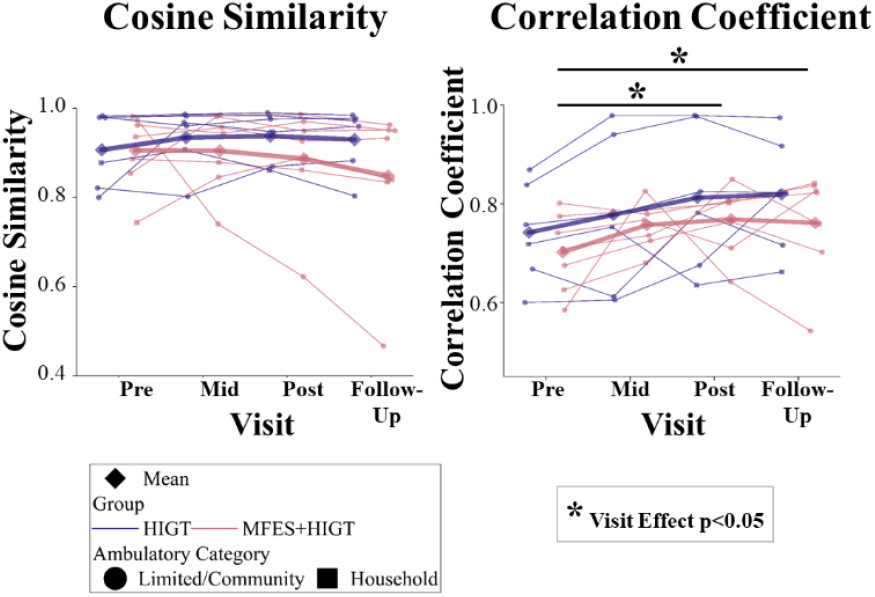
Muscle Synergy Metrics of Impaired Synergies at Baseline. Cosine similarity and circular cross correlation coefficient were computed between participant synergies that were impaired at baseline and average normative data. A value of one represents synergies that perfectly match the average normative data. Individual data points for each participant are displayed with household ambulators as squares and limited and community ambulators as circles. Significant visit effects are depicted by black lines. ^*^ (p < 0.05).

### D. Subgroup Analysis of Clinical Assessments

A clear limitation of this study was the ability of household ambulators to maintain target HR, spending an average of 22.9±14.3% in the target HR zone. This motivated investigation of the primary outcomes – clinical assessments – excluding more severely impaired participants (i.e., self-selected walking speed < 0.4 m/s, N=2 from MFES+HIGT, N=1 from HIGT). Results of this analysis are displayed in Fig. 6 and Supplemental Table II.

**Fig. 6:**
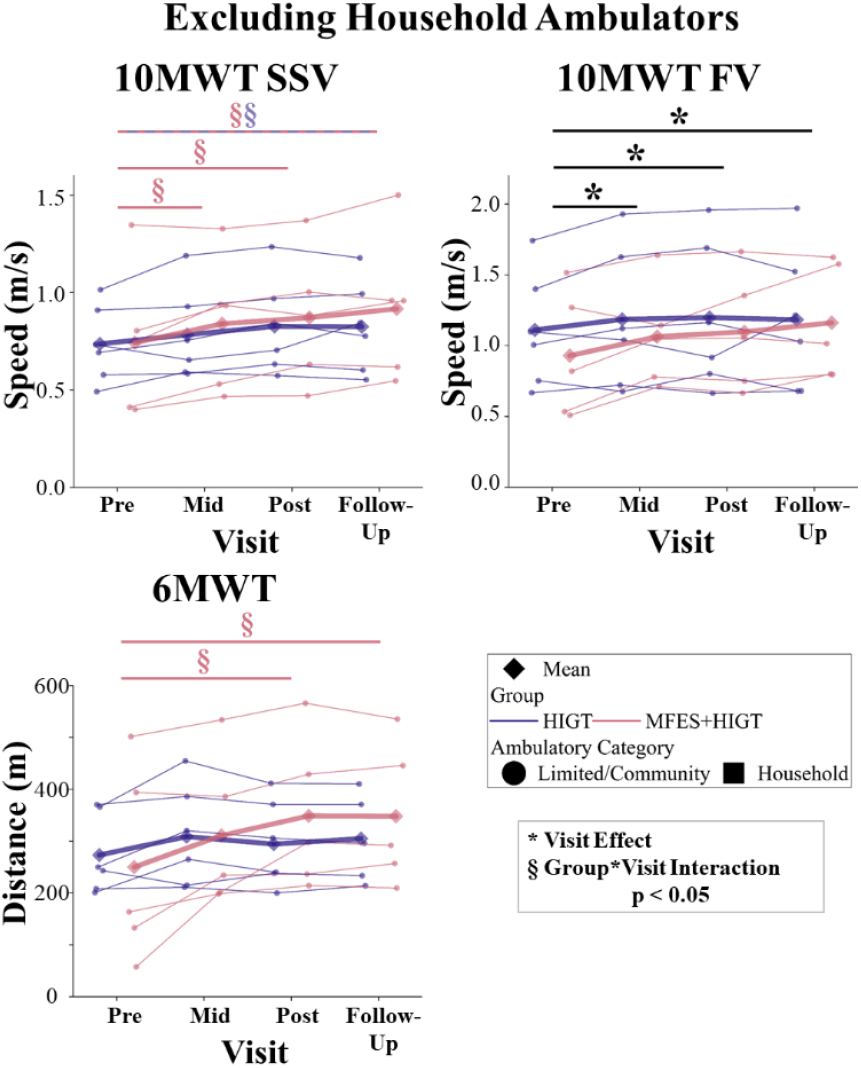
Clinical Assessments Excluding Household Ambulators. The 10 Meter Walk Test (10MWT) at self-selected velocity (SSV) and fast velocity (FV) and the 6 Minute Walk Test (6MWT) are presented. Individual data points for each participant are displayed, excluding household ambulators. Significant effects of visit are depicted by black lines and ^*^ where p<0.05. Significant group^*^visit interactions are depicted by lines matching the color of the group associated with the interaction and § where p<0.05.

10MWT SSV had an effect of visit (p<0.001) and an interaction (p=0.028), but no effect of group (p=0.849). Post-hoc analysis of the interaction revealed that the HIGT group only improved from pre to follow-up (p=0.017) while the MFES+HIGT group improved from pre to midpoint (p=0.028), post (p=0.003), and follow-up (p<0.001). The 10MWT FV had an effect of visit (p=0.009), but no effect of group (p=0.681) nor an interaction (p=0.122). Post-hoc analysis of visit revealed that both groups improved from pre to midpoint (p=0.029), post (p=0.011), and follow-up (p=0.042). Analysis of the 6MWT revealed an effect of visit (p=0.003) and an interaction (p=0.008), but no effect of group (p=0.542). Post-hoc analysis of the interaction revealed that only the MFES+HIGT group improved from pre to post (p<0.001) and follow-up (p=0.002). Average steps per day did not achieve significance (p>0.307).

## IV. Discussion

In this study, we investigated the feasibility of clinically implementing personalized synergy-based MFES with HIGT for chronic stroke gait rehabilitation and compared the efficacy with HIGT in a pilot study. Both groups were on average able to achieve target HR zones outlined for HIGT in clinical practice guidelines,[5] but household ambulators (N=3, baseline 10MWT SSV < 0.4 m/s) failed to maintain target HR. Physical therapists became proficient in preparing for MFES sessions quickly, plateauing in setup time around nine sessions at 4.53 minutes, which we believe to be feasible for clinical implementation. Additionally, therapists rated the feasibility, appropriateness, acceptability, and usability of the intervention highly. Despite the small sample size and non-significant but greater baseline function in the HIGT group, only the MFES+HIGT group demonstrated a significant improvement in endurance–although no difference between groups was detected at any time point. Both groups moderately improved walking speed, impaired limb step length, and similarity of synergy timings to controls with training. In an exploratory analysis, household ambulators were removed, as these participants did not achieve HIGT targets, and in this analysis, endurance and SSV improved only in the MFES+HIGT group from pre to midpoint and post.

### A. Training and Feasibility

On average, each group was able to achieve target HR zones (70-85%) for 50% of each session with no dropouts in the MFES+HIGT group (one dropout in the HIGT group due to a preexisting medical condition). This was consistent with prior studies,[7] where participants maintained target HR zones for at least 30% of a training session. However, the three household ambulators on average reached target HR zones 22.9±14.3% of the time. This is potentially due to the limited ability to increase speed and incline for these participants, thereby limiting our ability to effectively challenge them and elevate their heart rate. This is consistent with previous works where individuals with more severe gait impairments may not be able to tolerate or achieve target intensities during HIGT without assistance or technologies (e.g., exoskeleton).[5, 59] However, these participants did still rate the intervention as challenging using the RPE scale. Therefore, this intervention may be more appropriate for moderate and higher-functioning individuals.

The MFES+HIGT intervention was well-received and tolerated by both participants and therapists, with MFES+HIGT participants qualitatively reporting improved confidence, balance, and independence from assistive devices. After two training sessions led by the research team and nine independent sessions, therapists were able to prepare the intervention in 4.53 minutes on average. A study investigating currently used rehabilitation technologies in the clinical setting found that average setup time was 2.9±4.3 minutes, with 84% of technologies taking less than five minutes.[9] Therefore, this intervention appears to be feasible for clinical implementation, not taking away from the valuable time therapists have with patients. Additionally, the System Usability Scale indicated acceptable usability with an average of 71.5, passing the threshold for acceptable usability of 68/100, [57, 60] albeit in a small sample of four therapists. In addition, the Perceived Usefulness questionnaire indicated that therapists were likely to implement the intervention into their clinical practice. Therapists did address that, at this stage in the development of the device and the personalized protocol, they feel technical support is preferred. However, manual placement of the external hip unit led to greater setup times, lack of confidence when manually placing electrodes, and a few disconnects between the system and the external unit, which is when technical support was necessary. Further research is required to determine whether synergy-based MFES is feasible in a larger population with an improved device and intervention design.

### B. Training Efficacy

The present study found that while both groups demonstrated improvements in walking speed, impaired limb step length, and similarity of the activation profile of muscle synergies to normative data, only the MFES+HIGT group achieved significant and sustained gains in walking endurance, although no difference in improvement between groups was detected. Due to small sample size, we cannot conclude that MFES+HIGT promoted better improvements in endurance than HIGT, but these results support further investigation in a higher powered study to determine if synergy-based MFES outperforms HIGT. We did find that four out of seven participants in the MFES+HIGT group surpassed the MCID threshold for the 6MWT, compared to only one in the HIGT group. Endurance remains a critical challenge in stroke rehabilitation,[4] and greater walking endurance has been linked to better community reintegration outcomes.[61] Our findings are consistent with prior studies showing that extending FES to both the TA and gluteus medius can improve endurance in individuals with chronic stroke.[20, 21] Similarly, FastFES, which combines FES to the TA and GAS, has been shown to reduce the metabolic cost of walking.[22, 23]

We hypothesize that these endurance gains may be due to both peripheral [10] and central mechanisms.[11-15] Peripherally, FES may have improved propulsive force by enhancing activation of the gastrocnemius, a key muscle in the propulsive muscle synergy that was impaired in eleven out of fourteen participants at baseline. Although not directly measured, propulsion of the impaired limb is an essential predictor of 6MWT distance.[62] Repetitive, precisely timed FES may have increased muscle mass and strength,[63, 64] facilitating more efficient energy expenditure, as shown in previous FastFES studies.[22, 23] Centrally, we did observe that muscle synergy activation profiles became more similar to normative data in both groups and accompanied improved gait speed and endurance with training, which is consistent with previous work that has demonstrated that gait rehabilitation improves both synergies and gait outcomes.[29-31] Previous research has shown that FES can promote neuroplasticity and reshape neural coordination,[16, 17] and the muscle synergy-based stimulation technique has also been shown to reduce FES-induced fatigue in the upper limb compared to other stimulation strategies.[65] Biomechanical analysis also did not identify any differences between groups.

While spatial organization of synergies (i.e., cosine similarity) did not exhibit a significant change in either group, in the MFES+HIGT group, one participant dramatically reduced cosine similarity (no change in cross correlation coefficient). Notably, this participant achieved the greatest improvements in walking speed (SSV_Post-Pre_ = of 0.25 m/s) and distance (6MWT_Post-Pre_ = of 240 m). This participant also reduced hip abduction during swing by 5° (greatest among all participants) and increased hip extension at stance by 3.5° from pre to post, suggesting reduced compensatory movements and better propulsion. Therefore, the muscle synergy patterns for this participant were modified with training, but not in a way that we would expect, and it still resulted in a substantial functional improvement without negative biomechanical changes. As this is a single case, further research is required in a larger population to better characterize how muscle synergy patterns change as a result of synergy-based MFES training and investigate alternative changes in synergy organization that still minimize compensatory biomechanics.

Future research should also investigate different strategies to construct personalized MFES interventions based on an individual’s impaired muscle synergies, whether that be targeting the most impaired synergy, constructing based on the change in synergies that is required to replicate normative synergies, targeting both limbs as stroke indeed impairs both limbs (as was done in our pilot study [32]), or even providing feedback through a different modality, such as visual feedback. Furthermore, it is possible that with greater stimulation dosage and by accounting for the variability in baseline impairment, with a larger sample size, more substantial changes in functional outcomes, biomechanics, and synergy patterns with synergy-based MFES gait training may be better elucidated and responders and non-responders can be better characterized.

### C. Subgroup Analysis

Household ambulators did not achieve target HR zones and thus did not effectively receive the full benefits of either intervention. Therefore, we conducted an exploratory subgroup analysis where we investigated how the clinical assessments changed in mild-to-moderate impaired individuals (SSV>0.4 m/s [47]). Biomechanics and muscle synergy metrics were investigated, but no changes from the full dataset analysis were detected. The 6MWT still significantly improved for the statistical power required in randomized controlled trials. Broad inclusion criteria risk masking treatment effects due to high inter-individual variability, whereas overly restrictive criteria carry the risk of excluding those who may benefit. Therefore, pilot studies, such as the one presented here, are critical for refining inclusion and exclusion criteria, helping to identify the most appropriate target population for future randomized controlled trials. Ultimately, determining this balance may be the most important step toward advancing personalized, evidence-based interventions.

### D. Limitations

As previously mentioned, this study is limited by the small sample size of seven participants per group and four therapists, limiting the generalizability of these results. Furthermore, the baseline function, time since stroke, and age of the participants were quite variable with the HIGT group exhibiting slightly greater baseline function, although the difference was not significant. These limitations taken together potentially limited the statistical power and ability to achieve significant between group improvements. Furthermore, our preliminary finding of reduced recovery potential of household ambulators was limited to three participants and must be explored in a larger sample. This study also did not include blinded assessors, and only individuals in the MFES+HIGT group wore the sleeve during training, potentially leading to confounding effects of improvements due to compression. These limitations will need to be addressed in a future study evaluating the comparison of MFES+HIGT with HIGT in a randomized controlled trial. Furthermore, this study is limited to analysis of the nervous system at the periphery, but evidence has shown that FES can drive changes to neural coordination and activation at the MFES+HIGT from pre to post and follow-up while no changes were observed in the HIGT group.

Additionally, the subgroup analysis detected that only the MFES+HIGT group improved SSV from pre to midpoint and post, while both groups had improved from pre to follow-up. From these results, we cautiously conclude that synergy-based MFES+HIGT has the potential to improve both endurance and gait speed in mild-to-moderate impaired individuals. Furthermore, synergy-based MFES+HIGT may be most appropriate for mild and moderately impaired individuals because they can maintain target HR zones of HIGT to receive the full benefits of the intervention. One strategy to enhance the ability of more severely impaired individuals post-stroke to reach higher intensities is to combine synergy-based MFES with soft robotics in a hybrid solution, which may further enhance muscle coordination and joint kinematics with training.[66-68] We believe that this preliminary work supports further investigation into the feasibility in a larger population of therapists with greater diversity of experience as well as a randomized controlled trial to better characterize improvements achieved from synergy-based MFES and identify optimal responders and non-responders based on baseline function.

Our findings underscore a fundamental tension in clinical research: while personalization is essential for effective rehabilitation, it can also conflict with the central level.[11-15] Therefore, investigation of central mechanisms governing changes in muscle coordination will be essential to a deeper understanding of the mechanisms of recovery and could provide rationale behind responders and non-responders.

## V. Conclusion

In a small, variable sample of individuals living with lasting gait impairments due to chronic stroke, a personalized stimulation approach–muscle synergy-based multichannel functional electrical stimulation–paired with high-intensity gait training was feasibly implemented in the clinical setting and resulted in improved endurance and walking speed, particularly for participants with mild-to-moderate impairments. We posit that these preliminary results support pursuit of a larger, randomized controlled trial that can more effectively compare improvements between groups and identify responders based on baseline function. Leveraging approaches such as synergy-based MFES, which precisely tailors a rehabilitation intervention to an individual’s impaired neuromuscular coordination, holds great promise for translating personalized rehabilitation into clinical practice and enhancing recovery.

## Data Availability

Data will be available for reasonable request.

## References

[1] S. Strilciuc et al., “The economic burden of stroke: a systematic review of cost of illness studies,” (in eng), J Med Life, vol. 14, no. 5, pp. 606–619, 2021, doi: 10.25122/jml-2021-0361.

[2] C. L. Chen, H. C. Chen, S. F. Tang, C. Y. Wu, P. T. Cheng, and W. H. Hong, “Gait performance with compensatory adaptations in stroke patients with different degrees of motor recovery,” (in eng), Am J Phys Med Rehabil, vol. 82, no. 12, pp. 925–35, Dec 2003, doi: 10.1097/01.PHM.0000098040.13355.B5.

[3] G. Chen, C. Patten, D. H. Kothari, and F. E. Zajac, “Gait differences between individuals with post-stroke hemiparesis and non-disabled controls at matched speeds,” (in eng), Gait Posture, vol. 22, no. 1, pp. 51–6, Aug 2005, doi: 10.1016/j.gaitpost.2004.06.009.

[4] N. E. Mayo et al., “Disablement following stroke,” (in eng), Disabil Rehabil, vol. 21, no. 5-6, pp. 258–68, 1999, doi: 10.1080/096382899297684.

[5] T. G. Hornby et al., “Clinical Practice Guideline to Improve Locomotor Function Following Chronic Stroke, Incomplete Spinal Cord Injury, and Brain Injury,” Journal of Neurologic Physical Therapy, vol. 44, no. 1, pp. 49–100, 2020-01-01 2020, doi: 10.1097/npt.0000000000000303.

[6] A. Ojeda-Manzano, E. A. Pérez-Padilla, S. I. Sanguino-Suárez, C. Cabelka, H. Salgado, and A. Borstad, “The effects of high-intensity training on walking speed and endurance in the subacute phase poststroke: a systematic review,” Exploration of Neuroprotective Therapy, 2025-05-22 2025, doi: 10.37349/ent.2025.1004106.

[7] J. L. Moore et al., “Implementation of High-Intensity Stepping Training During Inpatient Stroke Rehabilitation Improves Functional Outcomes,” (in eng), Stroke, vol. 51, no. 2, pp. 563–570, 02 2020, doi: 10.1161/STROKEAHA.119.027450.

[8] C. Grefkes and G. R. Fink, “Recovery from stroke: current concepts and future perspectives,” Neurological Research and Practice, vol. 2, no. 1, 2020-12-01 2020, doi: 10.1186/s42466-020-00060-6.

[9] C. Celian et al., “Use of Technology in the Rehabilitation Setting: Therapy Observations, Mixed Methods Analysis, and Data Visualization,” Archives of rehabilitation research and clinical translation., vol. 7, no. 1, p. 100425, 2025, doi: 10.1016/j.arrct.2024.100425.

[10] P. E. Houghton et al., “Electrical stimulation therapy increases rate of healing of pressure ulcers in community-dwelling people with spinal cord injury,” (in eng), Arch Phys Med Rehabil, vol. 91, no. 5, pp. 669–78, May 2010, doi: 10.1016/j.apmr.2009.12.026.

[11] M. Gandolla, N. S. Ward, F. Molteni, E. Guanziroli, G. Ferrigno, and A. Pedrocchi, “The Neural Correlates of Long-Term Carryover following Functional Electrical Stimulation for Stroke,” (in eng), Neural Plast, vol. 2016, p. 4192718, 2016, doi: 10.1155/2016/4192718.

[12] M. Gandolla, L. Niero, F. Molteni, E. Guanziroli, N. S. Ward, and A. Pedrocchi, “Brain Plasticity Mechanisms Underlying Motor Control Reorganization: Pilot Longitudinal Study on Post-Stroke Subjects,” (in eng), Brain Sci, vol. 11, no. 3, Mar 05 2021, doi: 10.3390/brainsci11030329.

[13] G. I. Barsi, D. B. Popovic, I. M. Tarkka, T. Sinkjaer, and M. J. Grey, “Cortical excitability changes following grasping exercise augmented with electrical stimulation,” (in eng), Exp Brain Res, vol. 191, no. 1, pp. 57–66, Oct 2008, doi: 10.1007/s00221-008-1495-5.

[14] S. D. Iftime‐Nielsen, M. S. Christensen, R. J. Vingborg, T. Sinkjær, A. Roepstorff, and M. J. Grey, “Interaction of electrical stimulation and voluntary hand movement in SII and the cerebellum during simulated therapeutic functional electrical stimulation in healthy adults,” Human Brain Mapping, vol. 33, no. 1, pp. 40–49, 2012-01-01 2012, doi: 10.1002/hbm.21191.

[15] M. Milosevic, Y. Masugi, H. Obata, A. Sasaki, M. R. Popovic, and K. Nakazawa, “Short-term inhibition of spinal reflexes in multiple lower limb muscles after neuromuscular electrical stimulation of ankle plantar flexors,” (in eng), Exp Brain Res, vol. 237, no. 2, pp. 467–476, Feb 2019, doi: 10.1007/s00221-018-5437-6.

[16] C. Marquez-Chin and M. R. Popovic, “Functional electrical stimulation therapy for restoration of motor function after spinal cord injury and stroke: a review,” BioMedical Engineering OnLine, vol. 19, no. 1, 2020-12-01 2020, doi: 10.1186/s12938-020-00773-4.

[17] H. E. Shin et al., “Therapeutic Effects of Functional Electrical Stimulation on Physical Performance and Muscle Strength in Post-stroke Older Adults: A Review,” (in eng), Ann Geriatr Med Res, vol. 26, no. 1, pp. 16–24, Mar 2022, doi: 10.4235/agmr.22.0006.

[18] J. L. Allen, L. H. Ting, and T. M. Kesar, “Gait Rehabilitation Using Functional Electrical Stimulation Induces Changes in Ankle Muscle Coordination in Stroke Survivors: A Preliminary Study,” (in eng), Front Neurol, vol. 9, p. 1127, 2018, doi: 10.3389/fneur.2018.01127.

[19] S. Pereira, S. Mehta, A. McIntyre, L. Lobo, and R. W. Teasell, “Functional electrical stimulation for improving gait in persons with chronic stroke,” (in eng), Top Stroke Rehabil, vol. 19, no. 6, pp. 491–8, 2012, doi: 10.1310/tsr1906-491.

[20] M.-K. Cho, J.-H. Kim, Y. Chung, and S. Hwang, “Treadmill gait training combined with functional electrical stimulation on hip abductor and ankle dorsiflexor muscles for chronic hemiparesis,” Gait & posture., vol. 42, no. 1, pp. 73–78, 2015, doi: 10.1016/j.gaitpost.2015.04.009.

[21] Y. Chung, J.-H. Kim, Y. Cha, and S. Hwang, “Therapeutic effect of functional electrical stimulation-triggered gait training corresponding gait cycle for stroke,” Gait & posture., vol. 40, no. 3, pp. 471–475, 2014, doi: 10.1016/j.gaitpost.2014.06.002.

[22] L. N. Awad, D. S. Reisman, R. T. Pohlig, and S. A. Binder-Macleod, “Reducing The Cost of Transport and Increasing Walking Distance After Stroke: A Randomized Controlled Trial on Fast Locomotor Training Combined With Functional Electrical Stimulation,” (in eng), Neurorehabil Neural Repair, vol. 30, no. 7, pp. 661–70, Aug 2016, doi: 10.1177/1545968315619696.

[23] N. A. Hakansson, T. Kesar, D. Reisman, S. Binder-Macleod, and J. S. Higginson, “Effects of fast functional electrical stimulation gait training on mechanical recovery in poststroke gait,” (in eng), Artif Organs, vol. 35, no. 3, pp. 217–20, Mar 2011, doi: 10.1111/j.1525-1594.2011.01215.x.

[24] R. R. Neptune, D. J. Clark, and S. A. Kautz, “Modular control of human walking: A simulation study,” Journal of Biomechanics, vol. 42, no. 9, pp. 1282–1287, 2009-06-01 2009, doi: 10.1016/j.jbiomech.2009.03.009.

[25] H. Ting, Lena et al., “Neuromechanical Principles Underlying Movement Modularity and Their Implications for Rehabilitation,” Neuron, vol. 86, no. 1, pp. 38–54, 2015-04-01 2015, doi: 10.1016/j.neuron.2015.02.042.

[26] J. Gonzalez-Vargas, M. Sartori, S. Dosen, D. Torricelli, J. L. Pons, and D. Farina, “A predictive model of muscle excitations based on muscle modularity for a large repertoire of human locomotion conditions,” Frontiers in Computational Neuroscience, vol. 9, 2015-09-17 2015, doi: 10.3389/fncom.2015.00114.

[27] C. K. Balasubramanian, R. R. Neptune, and S. A. Kautz, “Variability in spatiotemporal step characteristics and its relationship to walking performance post-stroke,” (in eng), Gait Posture, vol. 29, no. 3, pp. 408–14, Apr 2009, doi: 10.1016/j.gaitpost.2008.10.061.

[28] D. J. Clark, L. H. Ting, F. E. Zajac, R. R. Neptune, and S. A. Kautz, “Merging of healthy motor modules predicts reduced locomotor performance and muscle coordination complexity post-stroke,” (in eng), J Neurophysiol, vol. 103, no. 2, pp. 844–57, Feb 2010, doi: 10.1152/jn.00825.2009.

[29] R. L. Routson, D. J. Clark, M. G. Bowden, S. A. Kautz, and R. R. Neptune, “The influence of locomotor rehabilitation on module quality and post-stroke hemiparetic walking performance,” Gait & Posture, vol. 38, no. 3, pp. 511–517, 2013-07-01 2013, doi: 10.1016/j.gaitpost.2013.01.020.

[30] C. K. Tan et al., “Corrigendum: Lateral Symmetry of Synergies in Lower Limb Muscles of Acute Poststroke Patients After Robotic Intervention,” (in eng), Front Neurosci, vol. 14, p. 113, 2020, doi: 10.3389/fnins.2020.00113.

[31] A. Cherubini, C. S. Del Valle, N. León, J. Tornero, J. A. Moreno, and C. B. Sanz-Morère, “Muscle Synergies During Gait of Functionally-Dependent Subacute Stroke Survivors Before and After Robotic Rehabilitation,” 2025 International Conference On Rehabilitation Robotics (ICORR), pp. 1059–1064, 2025, doi: 10.1109/ICORR66766.2025.11063159.

[32] S. Ferrante et al., “A Personalized Multi-Channel FES Controller Based on Muscle Synergies to Support Gait Rehabilitation after Stroke,” Frontiers neuroscience., vol. 10, 2016, 10.3389/fnins.2016.00425.

[33] J. Lim et al., “Patient-specific functional electrical stimulation strategy based on muscle synergy and walking posture analysis for gait rehabilitation of in doi: stroke patients,” Journal of International Medical Research, vol. 49, no. 5, p. 030006052110167, 2021-05-01 2021, doi: 10.1177/03000605211016782.

[34] Cionic. “Cionic Neural Sleeve.” https://cionic.com (accessed.

[35] U. S. F. a. D. Administration. “510(k) Premarket Notification: Cionic Neural Sleeve (K211651).” https://www.accessdata.fda.gov/scripts/cdrh/cfdocs/cfpmn/pmn.cfm?ID=K211651 (accessed.

[36] G. Borg, Borg’s Perceived Exertion and Pain Scales. Champaign, IL: Human Kinetics, 1998.

[37] J. W. Navalta et al., “Heart rate processing algorithms and exercise duration on reliability and validity decisions in biceps-worn Polar Verity Sense and OH1 wearables,” (in eng), Sci Rep, vol. 13, no. 1, p. 11736, Jul 20 2023, doi: 10.1038/s41598-023-38329-w.

[38] P. E. Oy. “Polar OH1 Optical Heart Rate Sensor.” Polar Electro Oy. https://www.polar.com/en/products/accessories/oh1-optical-heart-rate-sensor (accessed May 17, 2025).

[39] M. F. Rabbi, C. Pizzolato, D. G. Lloyd, C. P. Carty, D. Devaprakash, and L. E. Diamond, “Non-negative matrix factorisation is the most appropriate method for extraction of muscle synergies in walking and running,” Scientific Reports, vol. 10, no. 1, 2020-05-19 2020, doi: 10.1038/s41598-020-65257-w.

[40] S. J. Kamper, C. G. Maher, and G. Mackay, “Global Rating of Change Scales: A Review of Strengths and Weaknesses and Considerations for Design,” Journal of Manual & Manipulative Therapy, vol. 17, no. 3, pp. 163–170, 2009-07-01 2009, doi: 10.1179/jmt.2009.17.3.163.

[41] J. Brooke, “SUS—A Quick and Dirty Usability Scale,” Usability Evaluation in Industry, vol. 189, pp. 4–7, 1996.

[42] B. J. Weiner et al., “Psychometric assessment of three newly developed implementation outcome measures,” Implementation Science, vol. 12, no. 1, 2017-12-01 2017, doi: 10.1186/s13012-017-0635-3.

[43] A. Corp. “ActiGraph GT9X Link Activity Monitor.” ActiGraph Corp. https://www.actigraphcorp.com/ (accessed May 17, 2025).

[44] M. Ngueleu, C. Barthod, K. L. Best, F. Routhier, M. Otis, and C. S. Batcho, “Criterion validity of ActiGraph monitoring devices for step counting and distance measurement in adults and older adults: a systematic review,” (in eng), J Neuroeng Rehabil, vol. 19, no. 1, p. 112, Oct 17 2022, doi: 10.1186/s12984-022-01085-5.

[45] Q. Suau et al., “Current Knowledge about ActiGraph GT9X Link Activity Monitor Accuracy and Validity in Measuring Steps and Energy Expenditure: A Systematic Review,” (in eng), Sensors (Basel), vol. 24, no. 3, Jan 26 2024, doi: 10.3390/s24030825.

[46] J. R. Cotton et al., “Markerless Motion Capture and Biomechanical Analysis Pipeline,” arxiv, 2023, 10.48550/arXiv.2303.10654 <button aria-describedby=“more-info-desc-1” class=“more-info” style=“background-image: initial; backgroundposition: initial; background-size: initial; background-repeat: initial; background-attachment: initial; background-origin: initial; background-clip: initial; border-width: initial; border-style: none; border-color: initial; box-shadow: none; position: relative; padding: 0px 0px 0px 6px;”> Focus to learn more.

[47] M. G. Bowden, C. K. Balasubramanian, A. L. Behrman, and S. A. Kautz, “Validation of a speed-based classification system using quantitative measures of walking performance poststroke,” (in eng), Neurorehabil Neural Repair, vol. 22, no. 6, pp. 672–5, 2008, doi: 10.1177/1545968308318837.

[48] R. J. Cotton, “Differentiable Biomechanics Unlocks Opportunities for Markerless Motion Capture” arXiv preprint, 2024.

[49] “EasyMocap: Make human motion capture easier.” Github. https://github.com/zju3dv/EasyMocap (accessed.

[50] I Sárándi, A. Hermans, and B. Leibe, “Learning 3D Human Pose Estimation from Dozens of Datasets using a Geometry-Aware Autoencoder to Bridge Between Skeleton Formats,” in IEEE/CVF Winter Conference on Applications of Computer Vision (WACV), 2023: IEEE, pp. 297–306, doi: 10.1109/WACV56688.2023.00297. [Online]. Available: https://openaccess.thecvf.com/content/WACV2023/html/Sarandi_Learning_3D_Human_Pose_Estimation_From_Dozens_of_Datasets_Using_WACV_2023_paper.html

[51] R. J. Cotton, “PosePipe: Open-Source Human Pose Estimation Pipeline for Rehabilitation Research,” Archives of physical medicine and rehabilitation., vol. 103, no. 12, pp. e161–e162, 2022, doi: 10.1016/j.apmr.2022.08.868.

[52] R. J. Cotton, E. Mcclerklin, A. Cimorelli, A. Patel, and T. Karakostas, “Transforming Gait: Video-Based Spatiotemporal Gait Analysis,” in 2022 44th Annual International Conference of the IEEE Engineering in Medicine & Biology Society (EMBC), 2022-07-11 2022: IEEE, pp. 115–120, doi: 10.1109/embc48229.2022.9871036.

[53] A. Cimorelli, A. Patel, T. Karakostas, and R. J. Cotton, “Validation of portable in-clinic video-based gait analysis for prosthesis users,” Scientific Reports, vol. 14, no. 1, 2024-02-15 2024, doi: 10.1038/s41598-024-53217-7.

[54] M. C. Tresch, V. C. K. Cheung, and A. d’Avella, “Matrix Factorization Algorithms for the Identification of Muscle Synergies: Evaluation on Simulated and Experimental Data Sets,” Journal of neurophysiology /, vol. 95, no. 4, pp. 2199–2212, 2006, doi: 10.1152/jn.00222.2005.

[55] H.-Y. Kim, “Statistical notes for clinical researchers: assessing normal distribution (2) using skewness and kurtosis,” Restorative dentistry & endodontics., vol. 38, no. 1, p. 52, 2013, doi: 10.5395/rde.2013.38.1.52.

[56] S. G. West, J. F. Finch, and P. J. Curran, “Structural equation models with nonnormal variables: Problems and remedies,” in Structural Equation Modeling: Concepts, Issues, and Applications, R. H. Hyole Ed. Thousand Oaks, CA, US: Sage Publication, Inc 1995, pp. 56–75.

[57] A. Bangor, P. Kortum, and J. Miller, “Determining what individual SUS scores mean: Adding an adjective rating scale,” Journal of Usability Studies, vol. 4, no. 3, pp. 114–123, 2009.

[58] S. Perera, S. H. Mody, R. C. Woodman, and S. A. Studenski, “Meaningful Change and Responsiveness in Common Physical Performance Measures in Older Adults,” Journal of the American Geriatrics Society., vol. 54, no. 5, pp. 743–749, 2006, doi: 10.1111/j.1532-5415.2006.00701.x.

[59] E. C. Field-Fote and K. E. Roach, “Influence of a Locomotor Training Approach on Walking Speed and Distance in People With Chronic Spinal Cord Injury: A Randomized Clinical Trial,” Physical Therapy, vol. 91, no. 1, pp. 48–60, 2011-01-01 2011, doi: 10.2522/ptj.20090359.

[60] A. Bangor, P. T. Kortum, and J. T. Miller, “An Empirical Evaluation of the System Usability Scale,” International Journal of Human-Computer Interaction, vol. 24, no. 6, pp. 574–594, 2008-07-29 2008, doi: 10.1080/10447310802205776.

[61] M. Y. Pang, J. J. Eng, and W. C. Miller, “Determinants of satisfaction with community reintegration in older adults with chronic stroke: role of balance self-efficacy,” (in eng), Phys Ther, vol. 87, no. 3, pp. 282–91, Mar 2007, doi: 10.2522/ptj.20060142.

[62] L. N. Awad, S. A. Binder-Macleod, R. T. Pohlig, and D. S. Reisman, “Paretic Propulsion and Trailing Limb Angle Are Key Determinants of Long-Distance Walking Function After Stroke,” Neurorehabilitation and Neural Repair, vol. 29, no. 6, pp. 499–508, 2015-07-01 2015, doi: 10.1177/1545968314554625.

[63] J. Sun, F. Yan, A. Liu, T. Liu, and H. Wang, “Electrical Stimulation of the Motor Cortex or Paretic Muscles Improves Strength Production in Stroke Patients: A Systematic Review and Meta‐Analysis,” PM & R : the journal of injury, function, and rehabilitation., vol. 13, no. 2, pp. 171–179, 2021, doi: 10.1002/pmrj.12399.

[64] M. Nozoe et al., “Efficacy of neuromuscular electrical stimulation for preventing quadriceps muscle wasting in patients with moderate or severe acute stroke: A pilot study,” (in eng), NeuroRehabilitation, vol. 41, no. 1, pp. 143–149, 2017, doi: 10.3233/NRE-171466.

[65] S. Bala, V. Y. Vishnu, and D. Joshi, “Muscle Synergy-Based Functional Electrical Stimulation Reduces Muscular Fatigue in Post-Stroke Patients: A Systematic Comparison,” (in eng), IEEE Trans Neural Syst Rehabil Eng, vol. 31, pp. 2858–2871, 2023, doi: 10.1109/TNSRE.2023.3290293.

[66] F. Dell’Eva, S. Dalla Gasperina, M. Gandolla, A. Pedrocchi, and E. Ambrosini, “A hybrid FES-motor cooperative control over a knee joint movement: A feasibility study” in International Functional Electrical Stimulation Society 2022, Rotterdam, Netherlands, 2022.

[67] E. Tricomi et al., “Soft robotic shorts improve outdoor walking efficiency in older adults,” Nature Machine Intelligence, vol. 6, no. 10, pp. 1145–1155, 2024-10-01 2024, doi: 10.1038/s42256-024-00894-8.

[68] E. Ambrosini et al., “A Multifaceted Hybrid ES-Robotic Device for Gait Training in Individuals with Neurological Disorders,” Springer Science and Business Media LLC, 2024-09-26, 2024.

