## Supplemental Table 1 and 2 for "Synergy-based functional electrical stimulation for gait rehabilitation in chronic stroke: a pilot study"

SUPPLEMENTAL TABLE I

Intervention Efficacy

| Outcome | Group | Time Point | | | | Linear Mixed Model P-Value | | |
| --- | --- | --- | --- | --- | --- | --- | --- | --- |
|  |  | Pre | Mid | Post | Follow-Up | Group | Visit | Group*Visit |
| Clinical | | | | | | | | |
| 10MWT SSV (m/s) ↑ | MFES+HIGT | 0.57±0.43 | 0.64±0.44 | 0.70±0.45 | 0.69±0.50 | 0.705 | 0.001* | 0.735 |
|  | HIGT | 0.66±0.26 | 0.71±0.30 | 0.75±0.31 | 0.74±0.32 |  |  |  |
| 10MWT FV (m/s) ↑ | MFES+HIGT | 0.71±0.53 | 0.81±0.53 | 0.84±0.56 | 0.87±0.61 | 0.392 | 0.021* | 0.638 |
|  | HIGT | 0.99±0.48 | 1.06±0.56 | 1.07±0.58 | 1.05±0.57 |  |  |  |
| 6MWT (m) ↑ | MFES+HIGT | 191.3±183.8 | 237.4±173.3 | 264.7±187.4 | 259.5±188.2 | 0.990 | 0.011* | 0.026* |
|  | HIGT | 245.4±101.2 | 277.0±122.5 | 264.6±109.6 | 269.1±116.0 |  |  |  |
| Average Steps Per Day ↑ | MFES+HIGT | 2777±2089 |  | 2901±2007 |  | 0.406 | 0.701 | 0.991 |
|  | HIGT | 2040±1586 |  | 1904±1037 |  |  |  |  |
| Biomechanics | | | | | | | | |
| Step Length (m) ↑ | MFES+HIGT | 0.42±0.21 | 0.43±0.20 | 0.45±0.20 | 0.46±0.21 | 0.434 | 0.007* | 0.922 |
|  | HIGT | 0.48±0.11 | 0.49±0.14 | 0.51±0.12 | 0.52±0.13 |  |  |  |
| Step Width (mm)↓ | MFES+HIGT | 0.25±0.04 | 0.23±0.04 | 0.23±0.04 | 0.22±0.05 | 0.414 | 0.033* | 0.110 |
|  | HIGT | 0.22±0.05 | 0.21±0.06 | 0.22±0.05 | 0.22±0.05 |  |  |  |
| Single Support (%) ↑ | MFES+HIGT | 28.8±5.2 | 28.5±4.9 | 29.0±4.4 | 31.8±5.6 | 0.379 | 0.212 | 0.561 |
|  | HIGT | 31.1±3.9 | 31.1±4.3 | 31.1±4.4 | 31.7±5.1 |  |  |  |
| Double Support (%) ↓ | MFES+HIGT | 40.3±9.1 | 39.9±8.8 | 37.9±8.3 | 39.7±10.5 | 0.371 | 0.121 | 0.099 |
|  | HIGT | 36.4±6.5 | 35.4±6.9 | 35.7±6.3 | 35.9±7.8 |  |  |  |
| Ankle Dorsiflexion in Swing (°) ↑ | MFES+HIGT | -8.6±3.6 | -8.8±4.6 | -10.2±3.4 | -9.2±3.1 | 0.117 | 0.055 | 0.377 |
|  | HIGT | -4.4±5.9 | -7.3±4.5 | -8.0±4.3 | -.6.0±2.5 |  |  |  |
| Knee Extension in Stance (°) ↑ | MFES+HIGT | -5.2±6.5 | -5.5±5.1 | -2.5±3.9 | -2.5±2.5 | 0.625 | 0.889 | 0.139 |
|  | HIGT | -3.7±4.6 | -3.0±4.4 | 2.3±3.7 | -2.8±5.1 |  |  |  |
| Hip Extension in Stance (°) ↓ | MFES+HIGT | -9.9±12.8 | -11.1±9.1 | -13.8±10.4 | -11.7±11.7 | 0.479 | 0.040* | 0.216 |
|  | HIGT | -14.2±4.9 | -15.1±7.3 | -14.4±7.3 | -16.2±7.2 |  |  |  |
| Hip Abduction in Swing (°) ↑ | MFES+HIGT | -10.4±4.5 | -9.2±2.7 | -9.5±3.9 | -9.2±3.8 | 0.509 | 0.237 | 0.948 |
|  | HIGT | -9.1±3.2 | -8.2±3.0 | -8.2±3.1 | -8.3±3.0 |  |  |  |
| Muscle Synergies | | | | | | | | |
| Cosine Similarity ↑ | MFES+HIGT | 0.91±0.09 | 0.90±0.09 | 0.90±0.12 | 0.85±0.18 | 0.253 | 0.244 | 0.427 |
|  | HIGT | 0.91±0.08 | 0.93±0.07 | 0.94±0.06 | 0.93±0.07 |  |  |  |
| Cross Corr. Coefficient ↑ | MFES+HIGT | 0.70±0.07 | 0.76±0.05 | 0.77±0.07 | 0.76±0.10 | 0.473 | 0.005* | 0.850 |
|  | HIGT | 0.74±0.10 | 0.78±0.16 | 0.81±0.15 | 0.82±0.12 |  |  |  |
| Number of Synergies ↑ | MFES+HIGT | 3.0±0.6 | 2.9±0.7 | 3.1±0.9 | 3.1±0.7 | 0.767 | 0.766 | 1.000 |
|  | HIGT | 3.3±0.8 | 3.1±0.9 | 3.7±0.5 | 3.7±0.8 |  |  |  |
| ** p<0.05 for visit effects. § p<0.05 for group*visit interactions. The arrows indicate the direction of improvement. 10MWT = 10 Meter Walk Test, SSV = Self-Selected Velocity, FV = Fast Velocity, 6MWT = 6 Minute Walk Test* | | | | | | | | |

SUPPLEMENTAL TABLE II

Sub-Group Analysis of Clinical Assessments

| Outcome | Group | Time Point | | | | Linear Mixed Model P-Value | | |
| --- | --- | --- | --- | --- | --- | --- | --- | --- |
|  |  | Pre | Mid | Post | Follow-Up | Group | Visit | Group*Visit |
| 10MWT SSV (m/s) ↑ | MFES+HIGT | 0.74±0.39 | 0.84±0.35 | 0.87±0.35 | 0.92±0.38 | 0.849 | <0.001* | 0.028 § |
|  | HIGT | 0.74±0.20 | 0.78±0.24 | 0.83±0.25 | 0.82±0.24 |  |  |  |
| 10MWT FV (m/s) ↑ | MFES+HIGT | 0.93±0.45 | 1.06±0.37 | 1.10±0.42 | 1.16±0.41 | 0.681 | 0.009* | 0.122 |
|  | HIGT | 1.11±0.40 | 1.19±0.50 | 1.20±0.52 | 1.18±0.50 |  |  |  |
| 6MWT (m) ↑ | MFES+HIGT | 250.0±188.6 | 310.8±146.6 | 348.8±147.4 | 348.0±137.2 | 0.542 | 0.003* | 0.008 § |
|  | HIGT | 273.2±76.1 | 308.8±97.6 | 294.4±83.4 | 304.9±85.0 |  |  |  |
| Average Steps Per Day ↑ | MFES+HIGT | 3409±2189 |  | 3343±1888 |  | 0.307 | 0.619 | 0.811 |
|  | HIGT | 2173±1735 |  | 2023±1083 |  |  |  |  |
| ** p<0.05 for visit effects. § p<0.05 for group*visit interactions. The arrows indicate the direction of improvement. 10MWT = 10 Meter Walk Test, SSV = Self-Selected Velocity, FV = Fast Velocity, 6MWT = 6 Minute Walk Test* | | | | | | | | |
